# Vaccine effectiveness of two and three doses of BNT162b2 and CoronaVac against COVID-19 in Hong Kong

**DOI:** 10.1101/2022.03.22.22272769

**Authors:** Martina E. McMenamin, Joshua Nealon, Yun Lin, Jessica Y. Wong, Justin K. Cheung, Eric H. Y. Lau, Peng Wu, Gabriel M. Leung, Benjamin J. Cowling

## Abstract

**Background:** Hong Kong maintained extremely low circulation of SARS-CoV-2 until a major community epidemic of Omicron BA.2 starting in January 2022. Both mRNA BNT162b2 (BioNTech/Fosun Pharma) and inactivated CoronaVac (Sinovac) vaccines are widely available, however coverage has remained low in older adults. Vaccine effectiveness in this predominantly infection-naïve population is unknown.

**Methods:** We used individual-level case data on mild/moderate, severe/fatal and fatal hospitalized COVID-19 from December 31, 2021 to March 8, 2022, along with census information and coverage data of BNT162b2 and CoronaVac. We used a negative binomial model, adjusting for age and calendar day to estimate vaccine effectiveness of one, two and three dose schedules of both vaccines, and relative effectiveness by number of doses and vaccine type.

**Findings:** A total of 12.7 million vaccine doses were administered in Hong Kong’s 7.3 million population, and we analyzed data from confirmed cases with mild/moderate (N=5,474), severe/fatal (N=5,294) and fatal (N=4,093) COVID-19. Two doses of either vaccine protected against severe disease and death, with higher effectiveness among adults _≥_60 years with BNT162b2 (VE: 88.2%, 95% confidence interval, CI: 84.4%, 91.1%) compared to CoronaVac (VE: 74.1%, 95% CI: 67.8%, 79.2%). Three doses of either vaccine offered very high levels of protection against severe outcomes (VE: 98.1%, 95% CI: 97.1%, 98.8%).

**Interpretation:** Third doses of either BNT162b2 or CoronaVac provide substantial additional protection against severe COVID-19 and should be prioritized, particularly in older adults who received CoronaVac primary schedules. Longer follow-up is needed to assess persistence of different vaccine platforms and schedules.

**Funding:** COVID-19 Vaccines Evaluation Program, Chinese Center for Disease Control and Prevention

## INTRODUCTION

Hong Kong Special Administrative Region of China (Hong Kong; population 7.3 million) has pursued a COVID-19 elimination strategy since January 2020 involving stringent social distancing measures, border entry restrictions, isolation of cases and quarantine of close contacts, and the use of personal protective measures.^1^ Consequently, the disease had been largely controlled through December 2021 with four previous epidemic waves resulting in a total of 12,606 cases (<2 per 1,000) and 207 deaths (<3 per 100,000). Since February 2021, both inactivated (Sinovac; CoronaVac) and mRNA (BioNTech/Fosun Pharma; BNT162b2) vaccines have been widely available with residents offered the choice of either. However, by January 2022, two-dose vaccine coverage had only reached 46% in older adults 70-79 years of age and 18% in those aged _≥_80 years.^2^

A major community epidemic of COVID-19 Omicron variant (B.1.1.529) lineage BA.2 began in early January 2022, resulting in 649,454 laboratory confirmed cases, 313,127 cases reported by rapid antigen tests and nearly 5,000 deaths to March 17, 2022.^2,3^ Vaccination coverage has since risen steadily but remains low in the most vulnerable, with two-dose coverage at 66% and 37% in 70-79 and _≥_80 year olds respectively as of March 17, 2022. Third vaccine doses were recommended first for priority groups and then for the general public on 1 January 2022, to be given six months after the second dose.^4,5^ Third-dose uptake has been highest in the 40-59y age group (46% as of March 17, 2022) and lower in older adults (30% in 70-79 year olds; 10% in those _≥_80). Efforts to increase vaccine uptake in older and high-risk groups are underway, including reducing the duration between first and second doses for care home residents, extending vaccination clinic operating hours and deployment of vaccine outreach teams to care homes, housing estates and to residents with limited mobility.^6,7^

International data has shown vaccination with BNT162b2 reduces the frequency of severe outcomes, and to a lesser extent, infection for variants circulating prior to Omicron.^8–15^ Waning of protection has been observed in multiple contexts, in particular against infection,^16–18^ and recent studies have provided early indications of reduced effectiveness of BNT162b2 against the Omicron variant.^19–21^ Evidence on vaccine performance against the more transmissible Omicron subvariant BA.2 remains very limited, as is data on the performance of the inactivated CoronaVac vaccine.^22^ Limited observational evidence suggests strong and durable protection against severe disease and death, with transient protection against milder symptomatic disease.^23–26^ With a largely infection-naïve population and two COVID-19 vaccines in widespread use, Hong Kong represents a unique environment for monitoring vaccine effectiveness (VE) against Omicron BA.2. In this study we estimated VE of one, two and three doses of BNT162b2 and CoronaVac, their relative effectiveness, and the additional protection offered by third doses against mild/moderate infections, severe/fatal disease and death.

## METHODS

### Study design and population

We assessed VE of the BNT162b2 and CoronaVac vaccines using an ecological study design, which has been previously employed to provide estimates of VE in Israel.^27^ The study population consisted of residents of Hong Kong aged 20 years and over, where the population with zero, one, two or three doses of either vaccine at risk at a given time was derived using detailed data from the vaccination programme and population census. Information on all laboratory-confirmed SARS-CoV-2 cases in Hong Kong from December 31, 2021 to March 8, 2022 was obtained from nationwide individual level surveillance data provided by the Centre for Health Protection and linked to clinical outcome data provided by the Hospital Authority.

### Ethical approval

This project received approval from the Institutional Review Board of the University of Hong Kong.

### Infections and outcomes

Extensive PCR testing for SARS-CoV-2 is conducted in public hospitals, community test centres and private laboratories in Hong Kong. Testing is free-of-charge or available at low cost, and required for those who exhibit COVID-19 like symptoms, or following contact tracing based on exposure history or residential location. Regular screening is also required of certain professions, in particular those working with older adults or vulnerable persons. Positive rapid test results have been recognised as confirmed infections since February 25, 2022 and included in official case counts from March 7, 2022. Data on all laboratory-confirmed cases between December 31, 2021 and March 8, 2022 were extracted and cases classified as ‘imported’, i.e. detected in on-arrival quarantine, were excluded due to their non-representative SARS-CoV-2 exposure and vaccination histories. Sequencing of a subset of cases each day indicates that fewer than 1% of cases and deaths during the fifth wave have occurred with the Delta variant, with the remaining infections attributed to the Omicron BA.2 lineage.

Hong Kong has an advanced public and private healthcare system whereby private clinics comprise most primary care and government hospitals provide approximately 90% of hospital medical services at very low cost to patients.^28^ Up until mid-February 2022, all laboratory-confirmed COVID-19 cases were admitted to hospitals for isolation and standardized clinical management, regardless of symptom presentation, with their hospitalization records stored in the data system managed by the Hospital Authority. After mid-February 2022, due to the large number of incident cases, hospitalisation was reserved for patients with more severe disease, and milder cases were required to isolate at dedicated government quarantine facilities or at home. In the Hospital Authority data system, records of patients’ test results, medication and condition changes were documented and integrated into a centralized database from which we extracted relevant information on those experiencing mild/moderate disease prior to February 16, 2022 and severe disease and death at any time. We excluded those with conflicting information in the database, i.e. persons with a worst recorded condition of ‘mild’ but also experiencing a fatal outcome within hospital. Severe disease was defined as any severe, critical or fatal COVID-19 case (definitions for each in Appendix).

### Population uptake of COVID-19 vaccines

Data on the estimated population size at the end of 2021 by age and sex were obtained from the Census and Statistics Department of the Hong Kong Special Administrative Region Government. Data on the number of persons vaccinated with either the BNT162b2 or CoronaVac vaccines in Hong Kong each day since February 22, 2021 are available in a national vaccination database provided by the Department for Health. Data on all vaccinations that had occurred up to March 8, 2022, including vaccinee age and the type and date of receipt of each dose of vaccine, were extracted on March 10, 2022. Vaccination information for all cases in the surveillance data was cross checked with Hospital Authority records and any cases with discrepancies were excluded.

Those who received vaccines other than BNT162b2 or CoronaVac, or who received a mixed primary series of one dose of BNT162b2 and one dose of CoronaVac, were excluded from the analysis. In addition, for the purposes of this analysis we also exclude those who switched vaccine platform after the second dose, that is, those who received two doses of CoronaVac and a third dose of BNT162b2 and those who have received a primary series of BNT162b2 and a third dose of CoronaVac. Cases with known prior COVID-19 infection were also excluded.

### Statistical analysis

Incidence rates were calculated according to the number of doses of COVID-19 vaccination received (none, one, two or three) for each age group (20-29, 30-39, 40-49, 50-59, 60-69, 70-79, _≥_80 years) and calendar day throughout the study period. Additional stratification by vaccine type was included to estimate VE for each vaccine type and relative VE (rVE) between two and three doses of each vaccine. Vaccination status was categorised according to the date of vaccination plus a 14-day lag for all doses, to allow for the delay in immune response to vaccination. Daily numbers of persons in each vaccination category were inferred from the uptake data assuming that individuals received the same vaccine for first and second dose (aligned with Hong Kong guidelines), and using aggregate data by age on vaccine switching for the third dose. The population at risk in each stratum was matched to the report date of cases, and cumulative numbers of previous SARS-CoV-2 infections within each group were removed from the population at risk at each time point. Incidence rate ratios (IRR) were estimated using a negative binomial rate model for the daily counts of cases adjusted for age group and calendar day including the logarithm of person-time as an offset term in the model to account for differing numbers at risk within each strata. VE was defined as (1-IRR)×100%.

## RESULTS

A total of 486,074 persons had confirmed SARS-CoV-2 infection during the study period from December 31, 2021 to March 8, 2022. The case data were linked to the Hospital Authority dataset to determine their clinical outcomes and those with complete age and vaccination records were extracted. Of these, 5,474 persons were recorded as having mild/moderate disease between December 31, 2021 and February 15, 2022. During the entire study period from December 31, 2021 to March 8, 2022, 5,294 persons with severe/fatal disease and 4,093 with fatal disease were included (Table 1).

**Table 1.** Descriptive characteristics of confirmed COVID-19 cases in Hong Kong classified as having mild, severe or fatal disease between 31 December 2021 and 8 March 2022.

|  | Mild/moderate disease<br>(N= 5474) | Severe/fatal disease<br>(N=5294) | Fatal disease<br>(N=4093) |
| --- | --- | --- | --- |
| **Age** |  |  |  |
| 20-49 years | 3144 | 101 | 39 |
| 50-69 years | 1602 | 784 | 488 |
| ≥70 years | 728 | 4408 | 3566 |
| **Sex** |  |  |  |
| Male | 2337 | 3245 | 2528 |
| Female | 3137 | 2049 | 1565 |
| **Vaccination status<sup>a</sup>** |  |  |  |
| No doses | 1300 | 4064 | 3277 |
| One dose |  |  |  |
| *BNT162b2* | 151 | 73 | 44 |
| *CoronaVac* | 226 | 532 | 374 |
| Two doses |  |  |  |
| *BNT162b2* | 2139 | 130 | 74 |
| *CoronaVac* | 1271 | 434 | 287 |
| Three doses |  |  |  |
| *BNT162b2* | 126 | 12 | 7 |
| *CoronaVac* | 210 | 14 | 7 |
| **Median (25<sup>th</sup>, 75<sup>th</sup> percentile) of days between last vaccine dose and positive SARS-CoV-2 test result<sup>b</sup>** |  |  |  |
| One dose |  |  |  |
| *BNT162b2* | 27 (22, 35) | 21 (18, 32) | 21 (18, 32) |
| *CoronaVac* | 29 (21, 35) | 24 (17, 38) | 24 (17, 39) |
| Two doses |  |  |  |
| *BNT162b2* | 182 (151, 217) | 161 (73, 207) | 171 (91, 213) |
| *CoronaVac* | 179 (146, 209) | 127 (51, 162) | 125 (51, 157) |
| Three doses |  |  |  |
| *BNT162b2* | 31 (20, 49) | 52 (38, 70) | 68 (49, 77) |
| *CoronaVac* | 39 (25, 66) | 45 (24, 100) | 64 (30, 100) |

<sup>a</sup>Number of doses plus 14-day lag
<sup>b</sup>Median time since vaccination among those where 14 days has passed since latest dose

Up to March 8, 2022, a total of 12.7 million vaccine doses had been administered in Hong Kong. Severe disease or death occurred a median of 161 (interquartile range, IQR: 73 to 207) days after the second vaccination in those vaccinated with two doses of BNT162b2, and 127 (IQR: 51 to 162) among those who received two doses of CoronaVac. Those experiencing severe and fatal outcomes after a third dose tested positive a median of 52 (IQR: 38 to 70) days and 45 (IQR: 24 to 100) days after vaccination with BNT162b2 and CoronaVac respectively. The distribution of mild cases according to age and vaccination status were similar to the population, with severe disease and death occurring predominantly in the unvaccinated older population (Figure 2).

**Figure 1.**
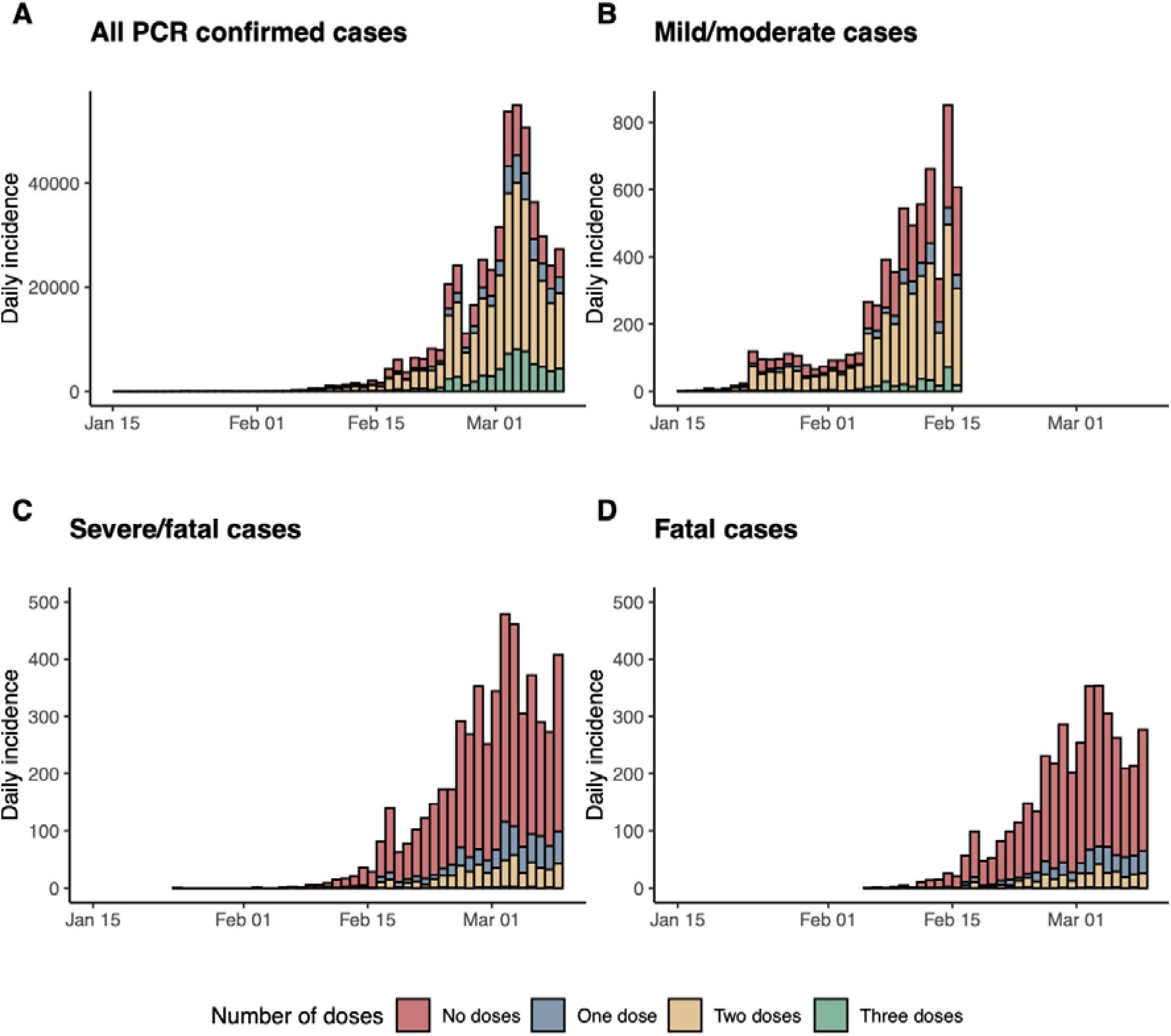
Daily incidence of (A) all PCR confirmed COVID-19 cases (B) mild/moderate cases in the early part of the fifth wave prior to 15 February 2022, (C) severe/fatal cases, and (D) deaths throughout the fifth wave in Hong Kong by vaccination status, where severe disease is defined as having ever been listed as ‘Serious’ or ‘Critical’ or ‘Fatal’ by the Hospital Authority during hospitalisation for COVID-19. Vaccination status was categorised according to the number of doses received plus a 14-day lag for all doses, to allow for the immune response to vaccination. The drop in mild/moderate cases on 4 March was due to a very small number of cases being reported as having been admitted to hospital or isolation facilities on that day. Mild cases were only included up until 15 February 2022 to account for change in admission criteria.

**Figure 2.**
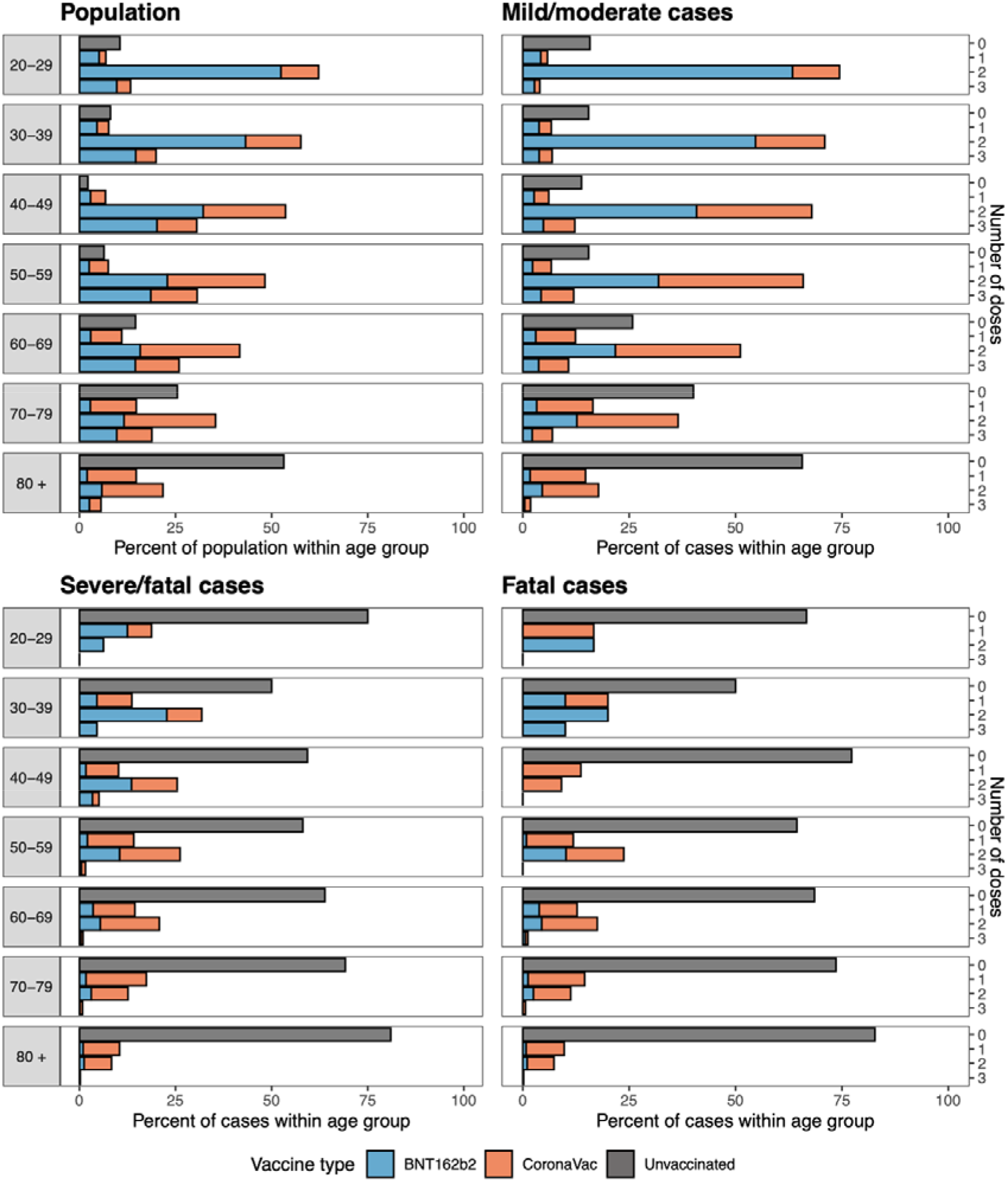
Vaccine status of population and those experiencing mild/moderate, severe/fatal and fatal COVID-19 as at 8 March 2022 as a percent of the population within a given age group shown by vaccine type and number of doses.

### VE after receipt of two doses

We found two doses of CoronaVac provided no protection against mild/moderate disease across all age groups, with some protection offered by BNT162b2 in younger age groups (VE: 31.0%, 95% CI: 1.6%, 51.7%). However, both vaccines were estimated to have high effectiveness against severe disease. Limited differences in vaccine effectiveness were observed for severe outcomes in younger adults, where VE was estimated to be 95.2% (95% CI: 92.9%, 96.8%) for BNT162b2 and 91.7% (95% CI: 87.8%, 94.4%) for CoronaVac (Table 2). The difference in VE was more pronounced for older adults, with higher effectiveness among adults >60 years who received BNT162b2 (VE: 88.2%, 95% confidence interval, CI: 84.4%, 91.1%) compared to CoronaVac (VE: 74.1%, 95% CI: 67.8%, 79.2%). When broken down further by age, we estimated that VE was 91.1% (95% CI: 85.4%, 94.6%) for BNT162b2 and 82.6% (74.2%, 88.2%) for CoronaVac in those 60-69y, reducing to 84.5% (95% CI: 75.5%, 90.2%) and 60.2% (95% CI: 43.9%, 71.8%) among those _≥_80y for BNT162b2 and CoronaVac, respectively. This was also observed for the mortality endpoint, where in adults aged _≥_80y two doses of BNT162b2 offered a higher level of protection against fatal disease (88.2%, 95% CI: 80.2%, 93.0%) compared to two doses of CoronaVac (66.8%, 95% CI: 51.9%, 77.0%).

**Table 2.**
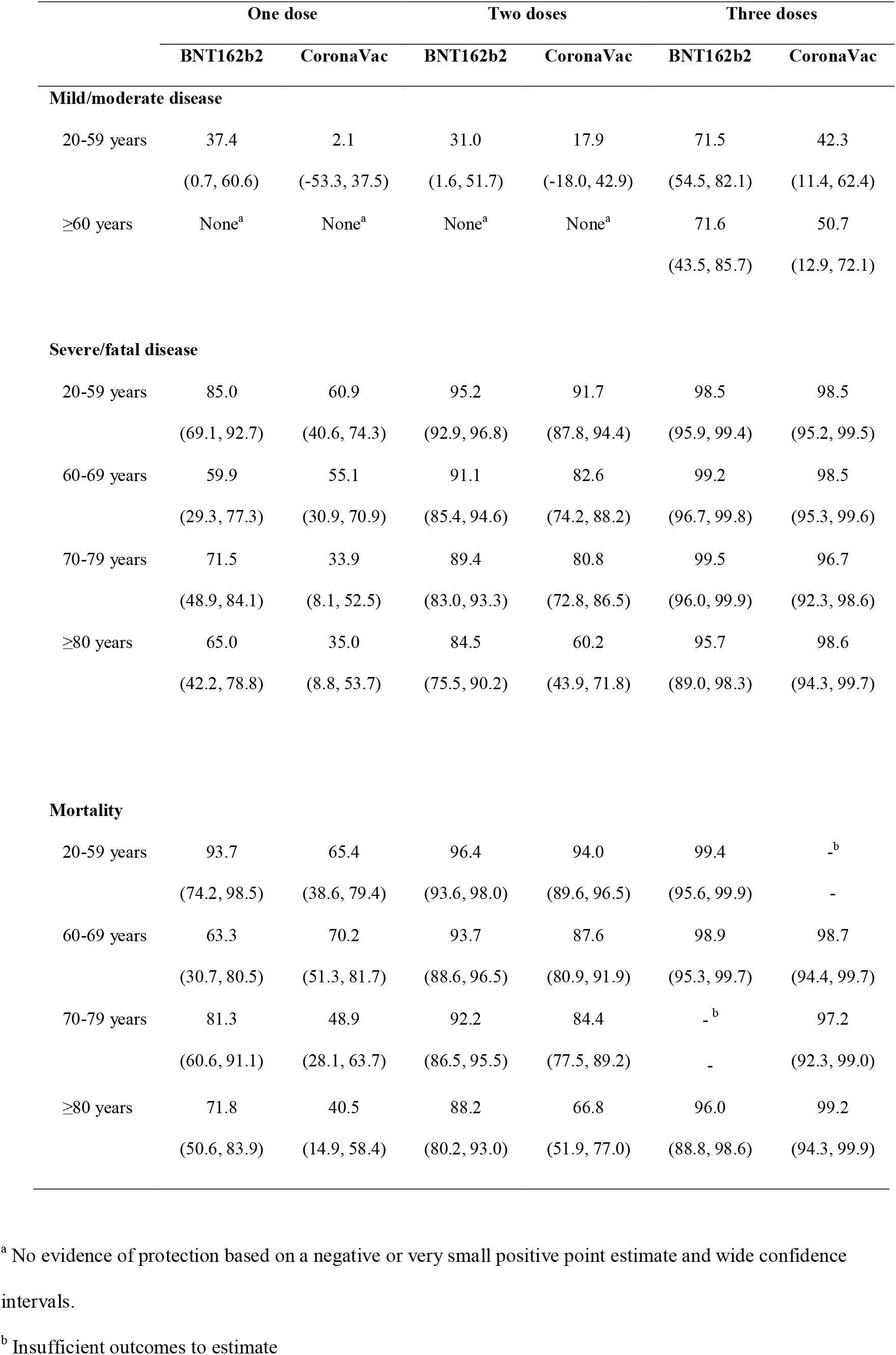
Vaccine effectiveness by dose (one, two, three) and vaccine type (CoronaVac, BNT162b2) in all ages and within age categories (mild/moderate: 20-59, _≥_60; severe/fatal, fatal: 20-59, 60-69, 70-79, _≥_80 years) against COVID-19 related mild/moderate disease, severe/fatal disease and death.

We compared the two-dose schedules of both vaccines and found no significant differences between BNT162b2 and CoronaVac for mild disease in any age group. Superiority of the two-dose BNT162b2 schedule was estimated for severe/fatal disease in adults _≥_60y (relative VE: 54.6%, 95% CI: 38.7%, 66.4%). This was also the case for mortality in those _≥_60y (relative VE: 58.5%, 95% CI: 70.7%, 41.3%). No differences between vaccines were found against severe/fatal or fatal COVID-19 in adults 20-59y.

### VE after receipt of three doses

We estimated three doses of both vaccines offered very high protection against severe disease (98.1%, 95% CI: 97.1%, 98.8%) and mortality (98.6%, 95% CI: 97.7%, 99.2%) which was sustained within all age groups (Table 2). Vaccine estimates were very similar for both vaccines against severe and fatal outcomes. Three doses of BNT162b2 was estimated to have a VE of 71.5% (95% CI: 54.5%, 82.1%) against mild/moderate disease in younger adults while for three doses of CoronaVac the VE was estimated as 42.3% (95% CI: 11.4%, 62.4%) against the same outcome.

### Relative VE of three versus two doses

We estimated the relative effect of three doses versus two doses of each vaccine type (Table 3). For mild/moderate disease we find an additional benefit of a third dose of BNT162b2 in younger (relative VE: 58.6%, 95% CI: 34.4%, 73.9%) and older (relative VE: 63.8%, 95% CI: 26.7%, 82.1%) adults who had previously received two doses of BNT162b2. A third dose of CoronaVac increased protection (relative VE: 57.0%, 95% CI: 23.4%, 75.9%) in older adults who had received two doses of CoronaVac, with no benefit observed in the younger age category. For severe/fatal disease we found an additional benefit of a third dose in adults of all ages for both vaccine types, with relative VE of 71.9% (95% CI: 25.1%, 89.5%) for three vs two doses of BNT162b2, and 96.6% (95% CI: 85.7%, 99.2%) for three vs two doses of CoronaVac among those _≥_80 years. Additional protection against mortality was offered by a third dose in older adults, with no differences observed in younger adults.

**Table 3.**
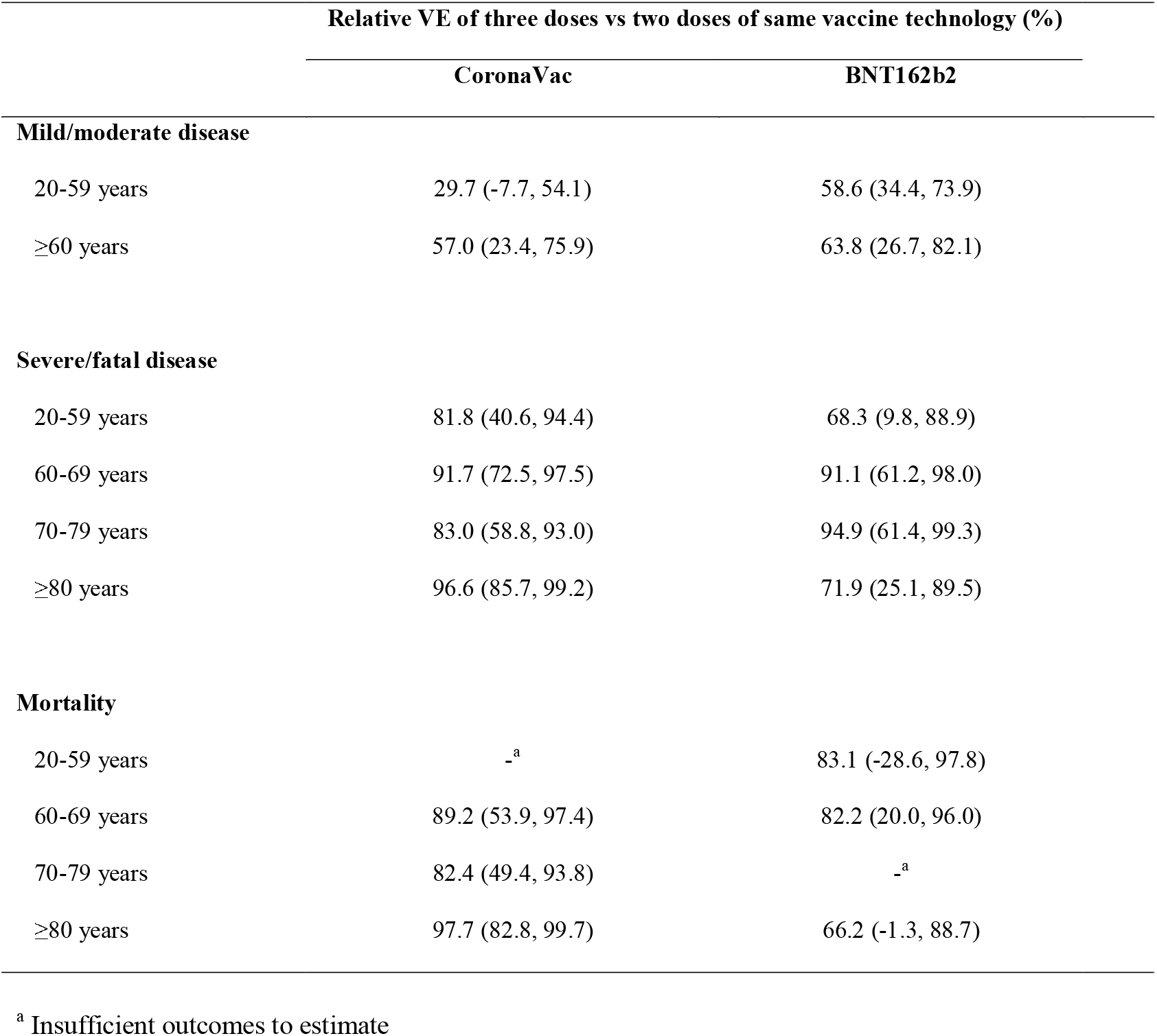
Relative vaccine effectiveness of a three versus two dose BNT162b2 schedule and a three versus two dose CoronaVac schedule against mild disease, severe disease and mortality as defined by the Hospital Authority.

|  | Relative VE of three doses vs two doses of same vaccine technology (%) |  |
| --- | --- | --- |
|  | CoronaVac | BNT162b2 |
| **Mild/moderate disease** |  |  |
| 20-59 years | 29.7 (-7.7, 54.1) | 58.6 (34.4, 73.9) |
| ≥60 years | 57.0 (23.4, 75.9) | 63.8 (26.7, 82.1) |
| **Severe/fatal disease** |  |  |
| 20-59 years | 81.8 (40.6, 94.4) | 68.3 (9.8, 88.9) |
| 60-69 years | 91.7 (72.5, 97.5) | 91.1 (61.2, 98.0) |
| 70-79 years | 83.0 (58.8, 93.0) | 94.9 (61.4, 99.3) |
| ≥80 years | 96.6 (85.7, 99.2) | 71.9 (25.1, 89.5) |
| **Mortality** |  |  |
| 20-59 years | -<sup>a</sup> | 83.1 (-28.6, 97.8) |
| 60-69 years | 89.2 (53.9, 97.4) | 82.2 (20.0, 96.0) |
| 70-79 years | 82.4 (49.4, 93.8) | -<sup>a</sup> |
| ≥80 years | 97.7 (82.8, 99.7) | 66.2 (-1.3, 88.7) |

<sup>a</sup> Insufficient outcomes to estimate

## DISCUSSION

We used detailed population-level data on the vaccination programme in Hong Kong since February 2021 and individual-level COVID-19 case data from December 31, 2021 to March 8, 2022 to estimate VE of one, two and three doses of BNT162b2 and CoronaVac vaccines in a largely infection-naïve population during the fifth wave of COVID-19 in Hong Kong. Two or three doses of BNT162b2 or three doses of CoronaVac provide a very high level of protection against severe disease and death in those under 80 years of age. A reduction in VE was observed among two-dose CoronaVac recipients _≥_80 years. We found no effect of two doses of CoronaVac and a limited effect of BNT162b2 against mild/moderate disease, with the caveat that many individuals had received their second dose several months before exposure to the SARS-CoV-2 virus. Limited protection against mild/moderate disease was restored with third doses for both vaccines, but we were only able to estimate VE for the short period since administration of third vaccine doses, and it is unclear how long this protection will last.

Although improved effectiveness of a third dose was observed against severe outcomes in younger age groups, the absolute VE of two doses remains high in this age group for both vaccines and the relative effects should be interpreted accordingly.^29^ Our finding that three doses of CoronaVac are needed for older adults to achieve high levels of protection is consistent with World Health Organization recommendations for this group.^30^ While there is a preferential recommendation in Hong Kong for a third dose of BNT162b2 in adults who received two doses of CoronaVac,^31^ this did not translate to preference in the community. Of all adults who had received two doses of CoronaVac and a third dose, only 26% received the third dose with BNT162b2. We were unable to evaluate the comparative effectiveness of heterologous vs homologous third dose schedules or durability of three dose protection in this study, but evidence from our analyses that three doses of inactivated vaccine provides a high level of protection against the severe spectrum of COVID-19 disease, at least in the short term, is reassuring.

Almost all sequenced SARS-CoV-2 isolates during Hong Kong’s fifth wave are of the Omicron BA.2 lineage. Our overall findings are largely consistent with existing VE evidence against this subvariant.^32–34^ A study from Qatar estimated that third dose VE for BNT162b2 was 43.7% (95% CI: 36.5, 50.0%) in the first month and begins to decline again in the following weeks, with substantially improved protection against severe outcomes (six-week VE: 90.9%, 95% CI: 78.6%, 96.1%).^35^ Similarly, a US study estimated VE of two doses of mRNA vaccines against severe Omicron disease, defined as COVID-19 requiring invasive mechanical ventilation or in-hospital death, of 79% (95% CI: 66%, 87%) a median of 265 days after the second dose; and three dose VE of 94% (95% CI: 88%, 97%), similar to our estimate of 98.1% (95% CI: 97.1%, 98.8%).^36^

Despite the overall consistency between our results and those presented in other studies, it is possible that VE, particularly against severe outcomes, has been overestimated in our study. Vaccine hesitancy in Hong Kong is highest among the elderly and in individuals with underlying health conditions.^37^ In this scenario so-called ‘healthy vaccinee bias’, by which vaccine recipients are healthier than their unvaccinated peers, may inflate the estimates.^38^ Although we have accounted for age in the current estimates, a lack of individual-level data on controls mean that this cannot be formally assessed with currently available data. However, our estimates for BNT162b2 and CoronaVac are similar to other studies using alternative designs, and we anticipate the magnitude of overestimation is unlikely to be substantial.^19,35^ Even if individual-level adjustments had been possible, estimating absolute VE after vaccines have been available for some time is problematic because it is necessary to compare incidence rates in vaccinated individuals with those from unvaccinated cohorts often with few remaining persons. This is the case in younger age groups in Hong Kong, whose characteristics are likely to differ substantially from those who chose to be vaccinated earlier. This bias, inherent to observational studies, is present in much of the existing VE literature at this stage of the pandemic. To address this concern, we also estimated a relative VE of three versus two doses of each vaccine type, as these cohorts are likely to be more comparable (Table 3). We find a third dose of either vaccine provides additional protection, reiterating the public health value of a third dose for minimizing severe disease and death but also for reducing health system congestion, public concern and indirect costs stemming from milder episodes during a COVID-19 epidemic.

We compared performance of the mRNA BNT162b2 and inactivated CoronaVac vaccines and found higher VE for BNT162b2 following one and two doses, but similar performance after three doses (Table 2). Our estimates are likely to be affected by time since vaccination, where typically more time has passed since administration of second than third doses which have only been widely available in Hong Kong since the beginning of January 2022 (Table 1). Improved effectiveness may partially reflect a recent, rather than a third, vaccine dose. This hypothesis is supported by data from an observational study in Malaysia which compared the duration of protection of the BNT162b2 and CoronaVac vaccines. They find more rapid waning of CoronaVac, in particular for mild/moderate and severe outcomes, but to a lesser extent for COVID-19 related mortality.^24^ Moreover, a recent study of humoral and cellular responses among Hong Kong vaccinees over time found that neutralising antibodies against variants of concern dropped to detection limit only three months after vaccinations, along with diminishing memory T cell responses, primarily among CoronaVac recipients.^39^

Our study has a number of limitations arising from available data and the nature of the epidemic within Hong Kong. Firstly, we used census data from the correct time period to construct the source population, but any differential population movement by vaccine status over the duration of the vaccination program could affect the validity of our estimates. Furthermore, as we are estimating vaccine effectiveness in real-time, there are large amounts of missingness in clinical data, which is especially problematic when assuming a population level denominator, as the assumed number of people still at risk will be overestimated. However, this is mostly an issue for mild/moderate outcomes, as we used complete records on COVID-19 mortality to derive estimates and we expect severe cases are fully documented. Secondly, there are some differences in testing requirements by vaccine status, particularly for those required to regularly test because of occupation. However, we expect that VE estimates against severe outcomes will be only marginally susceptible to biases related to testing requirements. Finally, in Hong Kong there was a clear preference for the BNT162b2 vaccine in younger age groups and for CoronaVac in older adults. We have addressed this confounding in estimates presented by stratifying by age categories and adjusting estimates by 10-year age categories and calendar day, however some residual confounding by age is possible in the vaccine platform-specific estimates and other factors may confound the relationship between vaccine status, type and risk of infection that cannot be accounted for in this design.

Our findings indicate that two dose schedules of both BNT162b2 and CoronaVac vaccines offer strong protection against severe disease and death, however higher levels of protection were observed among those who received two doses of BNT162b2 compared to those receiving two doses of CoronaVac, particularly in older age groups. Three recent doses of both vaccines offer very high levels of protection for older adults against severe outcomes, with no differences observed across vaccine types. It will be important to increase uptake of third vaccine doses, particularly in older adults who have so far received two doses of CoronaVac. Further investigation of the durability of protection provided by both vaccines is warranted and planned.

## Data Availability

All data produced in the present work are contained in the manuscript

## ACKNOWLEDGMENTS

The authors thank the Hong Kong Special Administrative Region Government and Hospital Authority for the timely sharing of COVID-19 vaccination and case data. The authors thank Julie Au for administrative support.

