## Appendix for "Vaccine effectiveness of two and three doses of BNT162b2 and CoronaVac against COVID-19 in Hong Kong"

**Supplementary Appendix**

Appendix Table 1 Definitions for severe and critical-to-fatal cases

| **Outcome** | **Classification** | **Criteria** |
| --- | --- | --- |
| Severe/fatal disease | Critical-to-fatal cases | Patient in Intensive Care Unit, or intubated, or require Extracorporeal Membrane Oxygenation (ECMO), or in shock |
|  |  | Fatal cases |
|  | Serious cases | Require oxygen supplement of 3 Litres per minute or more |
| Mild/moderate disease | Mild-to-moderate cases | Not being classified as either serious or critical as their worst condition during hospitalization, and not fatal. |

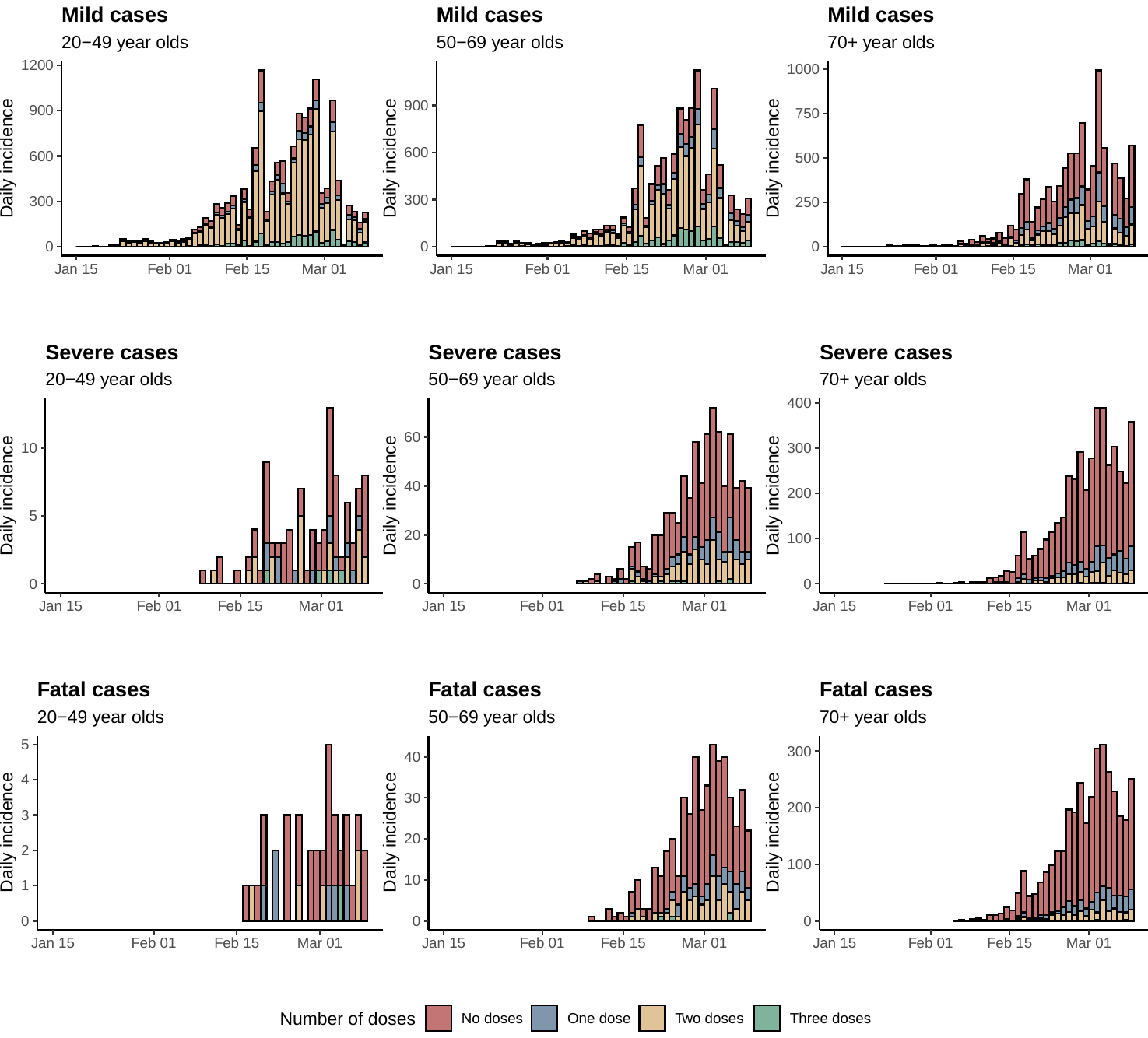

Appendix Figure 1 Daily incidence ‘mild’, ‘severe’ and fatal cases by age group and vaccination status, where mild was the worst recorded condition by HA and severe disease was defined as ever having been listed as ‘Serious’ or ‘Critical’ by HA during hospitalisation for COVID-19 or experiencing fatal COVID-19

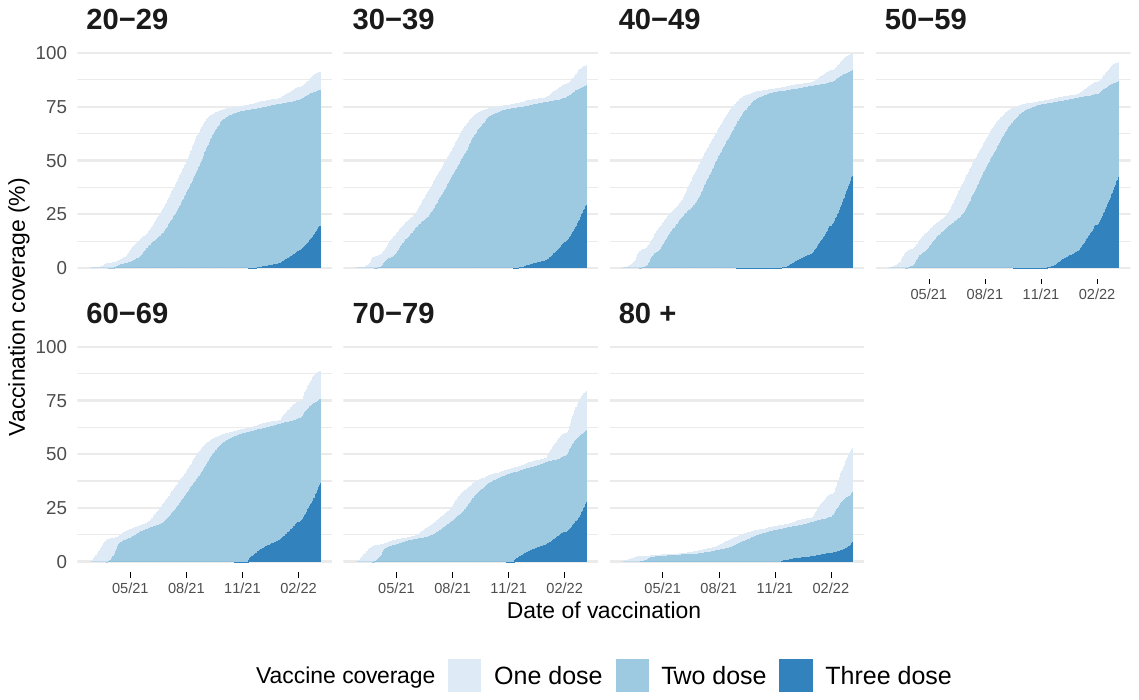

Appendix Figure 2 Vaccination coverage in Hong Kong for first, second and third doses of either BNT162b2 or CoronaVac vaccines by age group since the beginning of vaccination programme in Feb 2021

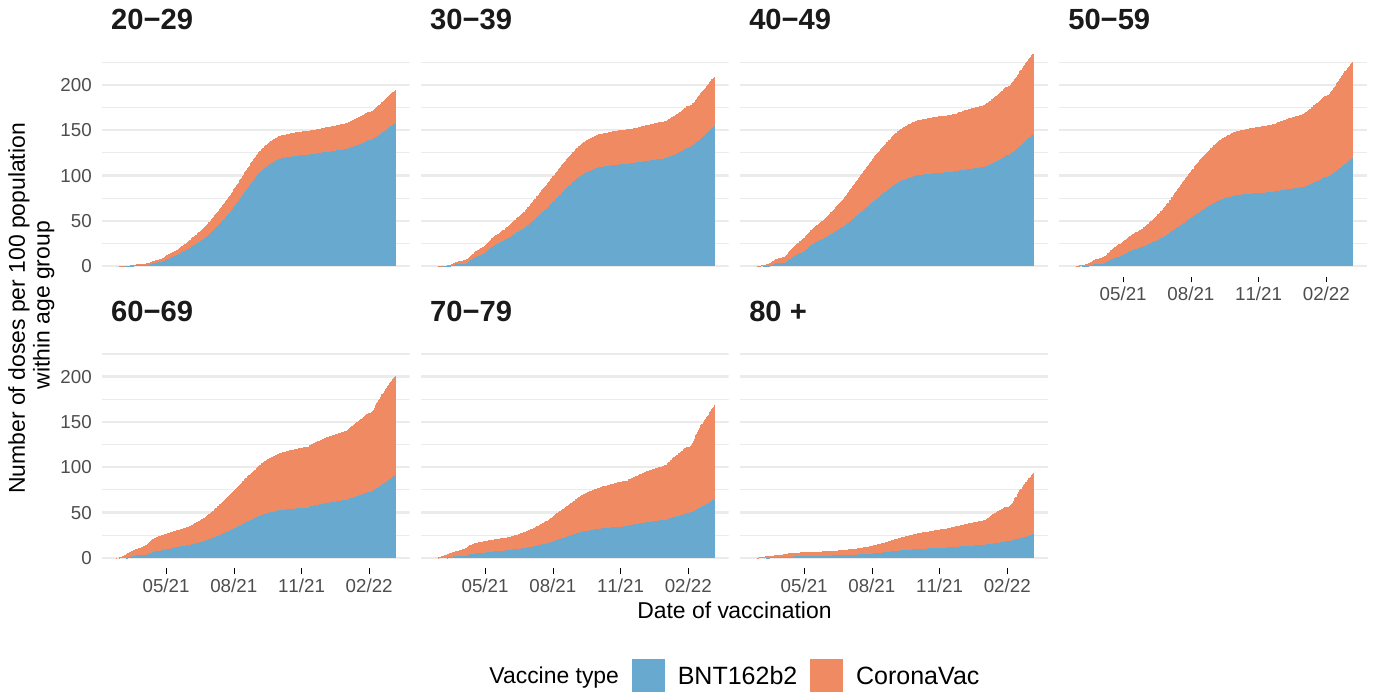

Appendix Figure 3 Number of doses administered over time throughout the vaccination programme in Hong Kong by vaccine type and age group. Note: Administration of CoronaVac vaccine started on February 26, 2021 and administration of BNT162b2 started on March 10, 2021

*Appendix Table 2. Overall absolute and relative vaccine effectiveness estimates for two or three dose schedules of either BNT162b2 and CoronaVac vaccines against mild/moderate infection, severe disease and death between 31 Dec 2021 and 08 Mar 2022 in Hong Kong, where estimates are obtained using a negative binomial rate model to estimate incidence rate ratio (IRR), and VE is defined as 1-IRR.*

|  | | | **Two doses** | | **Three doses** | |
| --- | --- | --- | --- | --- | --- | --- |
|  | | | **VE (%)** | | **VE (%)** | **rVE of three vs two doses** |
| **Mild/moderate disease** | | |  | |  |  |
| All ages |  | 6.6 (-21.6, 28.7) | | 50.9 (33.9, 63.7) | | 47.5 (31.9, 59.5) |
| 20-49 |  | 24.1 (-14.5, 49.7) | | 56.0 (30.5, 72.2) | | 42.1 (14.4, 60.8) |
| 50-69 |  | -10.3 (-25.5, 3.0) | | 44.0 (31.4, 54.2) | | 49.2 (39.3, 57.5) |
| 70 + |  | -12.0 (-89.4, 33.8) | | 67.4 (32.6, 84.2) | | 70.9 (42.9, 85.1) |
| **Severe/fatal disease** | |  | |  | |  |
| All ages |  | 85.0 (82.1, 87.5) | | 98.1 (97.1, 98.8) | | 87.2 (80.8, 91.8) |
| 20-49 |  | 95.4 (92.1, 97.4) | | 97.6 (93.2, 99.2) | | 47.9 (-54.4, 82.4) |
| 50-69 |  | 89.0 (85.6, 91.6) | | 98.8 (97.6, 99.4) | | 89.3 (77.8, 94.8) |
| 70 + |  | 76.4 (69.8, 81.5) | | 97.4 (95.4, 98.5) | | 89.0 (80.5, 93.8) |
| **Mortality** | |  | |  | |  |
| All ages |  | 87.4 (84.6, 89.7) | | 98.6 (97.7, 99.2) | | 89.0 (81.4, 94.0) |
| 20-49 |  | 97.9 (94.4, 99.2) | | 98.8 (90.8, 99.8) | | 41.4 (-404.8, 93.2) |
| 50-69 |  | 91.8 (88.8, 94.0) | | 99.2 (97.8, 99.7) | | 90.1 (72.7, 96.4) |
| 70 + |  | 80.6 (74.8, 85.0) | | 98.0 (96.0, 99.0) | | 89.6 (79.2, 94.8) |

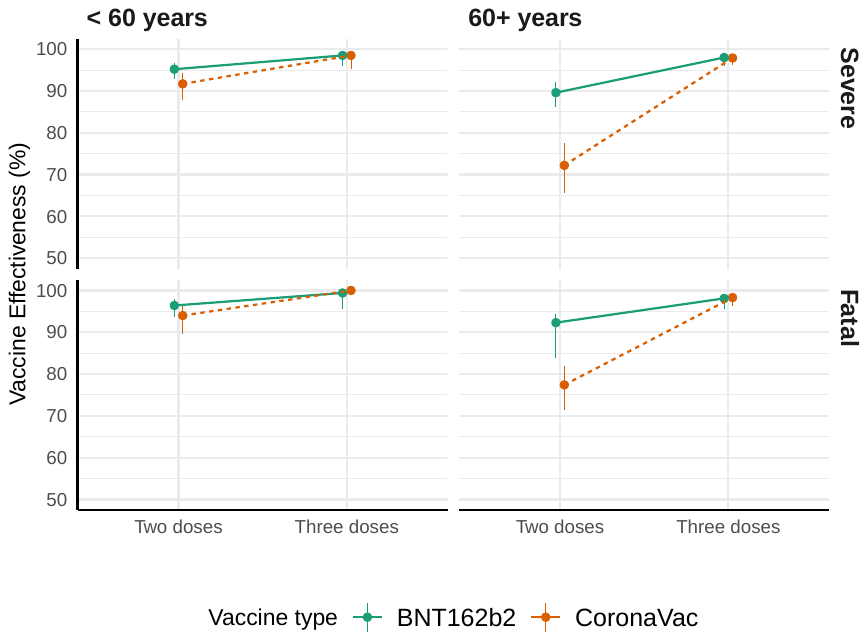

Appendix Figure 4 Vaccine effectiveness of two and three dose schedules for BNT162b2 and CoronaVac vaccines in those aged under 60 and 60 + against severe and fatal COVID-19
